# SARS-CoV-2 spread across the Colombian-Venezuelan border

**DOI:** 10.1101/2020.07.09.20149856

**Authors:** Alberto Paniz-Mondolfi, Marina Muñoz, Carolina Florez, Sergio Gomez, Angelica Rico, Lisseth Pardo, Esther C. Barros, Carolina Hernández, Lourdes Delgado, Jesús E. Jaimes, Luis Pérez, Aníbal A. Teherán, Hala Alejel Alshammary, Ajay Obla, Zenab Khan, Ana S. Gonzalez-Reiche, Matthew M. Hernandez, Emilia Mia Sordillo, Viviana Simon, Harm van Bakel, Juan David Ramírez

## Abstract

**Introduction:** Venezuela and Colombia both adopted measures of containment early in response to the COVID-19 pandemic. However, Venezuela’s ongoing humanitarian crisis has decimated its health care system, and forced millions of Venezuelans to flee through its porous border with Colombia. The extensive shared border, and illegal cross-border transit through improvised trails between the two countries are major challenges for public health authorities. We report the first SARS-CoV-2 genomes from Venezuela, and present a snapshot of the SARS-CoV-2 epidemiologic landscape in the Colombian-Venezuelan border region.

**Methods:** We sequenced and assembled viral genomes from total RNA extracted from nasopharyngeal (NP) clinical specimens using a custom reference-based analysis pipeline. Three assemblies obtained were subjected to typing using the Phylogenetic Assignment of Named Global Outbreak LINeages ‘Pangolin’ tool. A total of 376 publicly available SARS-CoV-2 genomes from South America were obtained from the GISAID database to perform comparative genomic analyses. Additionally, the Wuhan-1 strain was used as reference.

**Results:** We found that two of the SARS-CoV-2 genomes from Venezuela belonged to the B1 lineage, and the third to the B.1.13 lineage. We observed a point mutation in the Spike protein gene (D614G substitution), previously reported to be associated with increased infectivity, in all three Venezuelan genomes. An additional three mutations (R203K/G204R substitution) were present in the nucleocapsid (N) gene of one Venezuelan genome.

**Conclusions:** Genomic sequencing demonstrates similarity between SARS-CoV-2 lineages from Venezuela and viruses collected from patients in bordering areas in Colombia and from Brazil, consistent with cross-border transit despite administrative measures including lockdowns. The presence of mutations associated with increased infectivity in the 3 Venezuelan genomes we report and Colombian SARS-CoV-2 genomes from neighboring borders areas may pose additional challenges for control of SARS-CoV-2 spread in the complex epidemiological landscape in Latin American countries. Public health authorities should carefully follow the progress of the pandemic and its impact on displaced populations within the region.

## 1. INTRODUCTION

As Severe Acute Respiratory Syndrome Coronavirus −2 (SARS-CoV-2) spreads throughout the Western hemisphere, Latin America has become an epicenter for the Coronavirus Disease 2019 (COVID-19) pandemic. Since the report on March 13, 2020 of the first two cases diagnosed with COVID-19 in Venezuela, SARS-CoV-2 has spread rapidly across the country. The areas of highest transmission flank the Colombian-Venezuelan border, where the reported incidence reaches 47,9% with a 34,2% case fatality rate (“Coronavirus Venezuela,” n.d.)

Venezuela’s ongoing humanitarian crisis has had a severe impact on its health care system: the country has experienced a massive exodus of medical personnel, shortages of treatment and supplies, as well as a systematic dismantling of public health infrastructure (Daniels, 2020). In addition, the increased poverty and violence, plus the abandonment of all epidemiological surveillance programs, has set the stage for the re-emergence of vaccine-preventable and vector-borne diseases (Grillet et al., 2019; Paniz-Mondolfi et al., 2019). The Venezuelan public health system cannot withstand current autochthonous threats, let alone emerging infectious agents such as SARS-CoV-2. Furthermore, the political and economic turmoil in Venezuela has precipitated one of the largest refugee crises witnessed in the hemisphere. This has resulted in the establishment of disease corridors to neighboring countries---particularly Colombia, which houses 1.4 million Venezuelan migrants (Daniels, 2020; Torres and Castro, 2019). Despite early implementation of lockdown and border restrictions starting in March 2020, control of cross-border migration has remained a challenge due to the length (approximately 2,219 km) of the Venezuelan-Colombian border, and the unregulated transit of individuals through illegal trails known as “trochas”. As of July 8^th^ 2020, the majority of COVID-19 cases in Venezuela have been reported from three border states, Apure, Táchira and Zulia. (“Estadísticas Venezuela | COVID-19 en Venezuela,” n.d.).

Here we report sequences for three of five SARS-CoV-2-positive samples, representing the first viral genomes from Venezuela, and providing a snapshot of the epidemiological landscape across the Colombian-Venezuelan border.

## 2. METHODS

### 2.1. Patients, sampling and demographic data

Newly arrived migrants from Venezuela meeting case-definition criteria established by the Colombian Ministry of Health and Social Protection were screened for SARS-CoV-2 infection at different hospitals and healthcare centers in Norte de Santander and Bolivar Departments of Colombia between March 31^st^ and May 1^st^, 2020. Molecular detection of SARS-CoV-2 in nasopharyngeal swab specimens in viral transport media (NP-VTM) was performed using the Berlin Charité protocol. SARS-CoV-2 -positive specimens from five individuals who were tested within 24 hours of their arrival in Colombia were referred for further characterization. Four of the five individuals already were symptomatic at the time of arrival from Venezuela, and one was a close contact of a confirmed COVID-19 patient. Complete viral genomes were generated from specimens from three of these five cases. The patients, aged 30 (♂), 42 (♂) and 56 (♀) years, came from different regions, Cucuta, Norte de Santander, and Bolivar, (Fig 1A). Sequencing was unsuccessful for the other 2 specimens, most likely due to insufficient target material.

**Figure 1.**
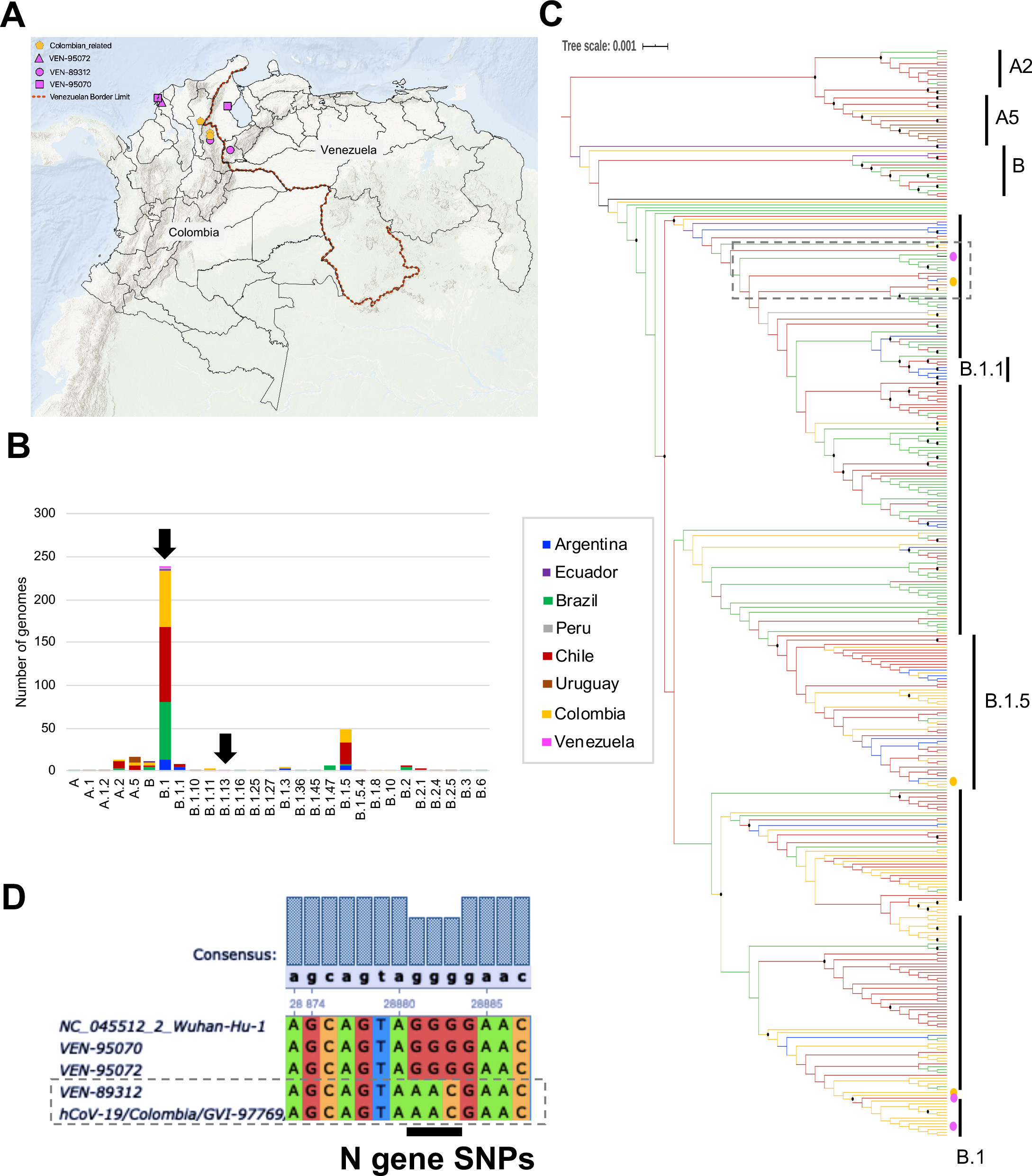
Regional comparative genomic analysis of SARS-CoV-2. **A**. Geographical distribution of the sequences from Venezuelan genomes and the available Colombian genomes analyzed in this study. The pink symbols indicate the Venezuelan patients identified in Colombia with their respective Venezuelan state origin. **B**. Stacked bar plot of the number of genomes per lineage determined using Phylogenetic Assignment of Named Global Outbreak LINeages ‘Pangolin’ tool. Three SARS-CoV-2 genomes from Venezuela were compared with 376 assemblies from other 7 South American countries (Argentina, Brazil, Chile, Colombia, Ecuador, Peru and Uruguay) using the publicly available GISAID EpiCoV™ database (https://www.gisaid.org/). Frequencies are discriminated by country of origin. The Venezuelan lineages are identified by black arrows. **C**. Maximum likelihood tree built in IQtree shows the phylogenetic relationships between genomes from Venezuela (pink dots) and the closest Colombian regions (yellow dots) with other South American genomes. The branches were colored according to the country of origin, using the color code of A panel. The clustering of the most frequent pangolin lineages (n> 10) is represented on the right side. The black dots represent highly supported nodes. **D**. Multiple alignment of the Nucleocapsid gene showing the substitutions found using the Wuhan-1 sequence as reference.

### 2.2. Phylogenetic analysis

We sequenced and assembled viral genomes from total RNA extracted from NP viral transport medium (VTM) clinical specimens, as described previously (Gonzalez-Reiche et al., 2020; Ramirez et al., 2020). The data generated by high throughput sequencing was assembled using a custom reference-based analysis pipeline (Gonzalez-Reiche et al., 2020). The complete genomes were typed using the Phylogenetic Assignment of Named Global Outbreak LINeages ‘Pangolin’ tool (Rambaut et al., 2020).

A total of 376 publicly available SARS-CoV-2 genomes encompassing the lineage diversity from South America were downloaded from the GISAID database for comparative genomic analyses. These sequences were aligned in MAFFT (Katoh et al., 2018), using the Wuhan-1 strain (NC_045512.2) as reference. The complete data set was subjected to the same typing scheme using Pangolin tool. Thus far, 28 pangolin lineages have been reported for SARS-CoV2 strains circulating in South American countries (Fig. 1B), with B1 as the predominant lineage representing 62.9% of the total reported genomes. Four other lineages include B.1.5 (12.5%), A.5 (4.5%), A.2 and B (3.4% each) account for an additional 23.8%. Each of the remaining 15 lineages accounted for fewer than 10 genomes, with several having only a single representative. A time-scaled maximum likelihood (ML) phylogeny based on TreeTime built in IQTREE (Rambaut et al., 2016; Sagulenko et al., 2018) revealed that there was no clustering by originating country in the reconstruction, although a general clustering by pangolin lineages was observed (Fig. 1C). These analyses are in agreement with the simultaneous circulation of SARS-CoV-2 lineages from different geographical origins.

A detailed screening of single-nucleotide polymorphisms (SNPs) in important open reading frames (ORFs) of SARS-CoV-2 was then conducted, and substitutions in Spike and Nucleocapsid sequences were evaluated. The alignment was inspected using Ugene (http://ugene.net/). The regions of interest were exported considering the ORFs described for the reference strain Wuhan-1 (NC_045512.2), as previously described in NCBI: https://www.ncbi.nlm.nih.gov/nuccore/?term = Severe + acute + respiratory + syndrome + coronavirus + 2 + isolate + Wuhan-Hu-1.

## 3. RESULTS

We found that two of the SARS-CoV-2 genomes obtained (VEN-89312 and VEN-95072) were identical and belonged to the B1 lineage, while the third (VEN-95070) belonged to the B.1.13 lineage, suggesting two separate introductions (Candido et al., 2020; Ramirez et al., 2020). We analyzed the individual genomic information for the 3 Venezuelan viruses reported in this study within the context of the larger global viral diversity to determine potential dates for the introduction of SARS-CoV2 into Venezuela, and similarly for 3 viruses collected from patients in bordering areas in Colombia (Ramirez et al., 2020) (Table 1).

**Table 1.**
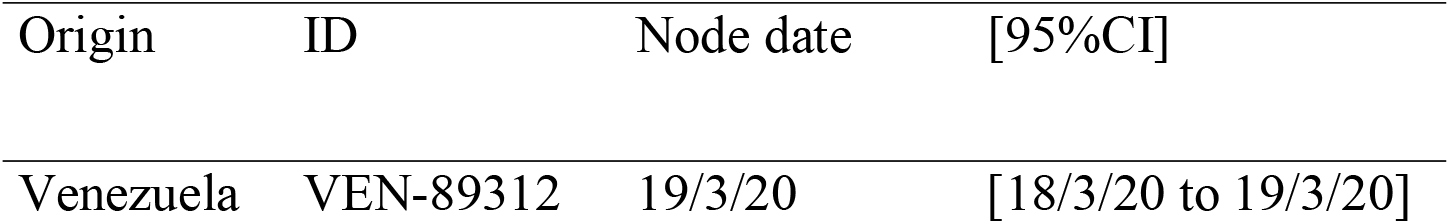
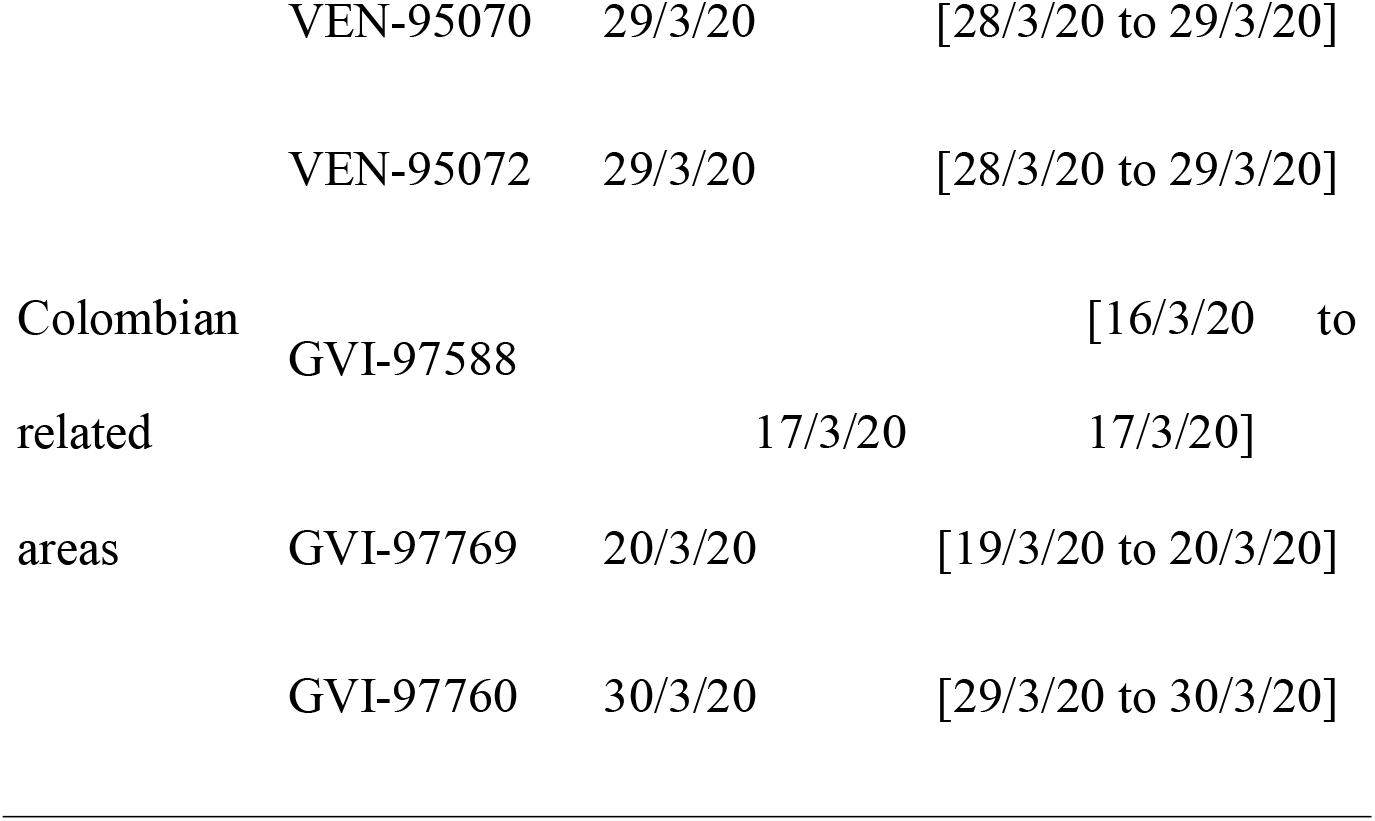
Potential introduction dates of SARS-CoV-2 from Venezuelan patients and patients in related border areas in Colombia.

Phylogenetic analysis revealed that two of the Venezuelan SARS-CoV-2 genomes (VEN-95070 and VEN-95072) closely resembled genomes from neighboring Colombia (Cesar and Norte de Santander Departments) (Ramirez et al., 2020) while the third (VEN-89312) was related to genomes from Brazil (Fig. 1C) (Candido et al., 2020).

The three Venezuelan genomes carried a G-to-A point mutation at position 23,403 resulting in a D614G substitution in the spike (S) protein. This mutation characterizes the B.1 lineages and has been associated with enhanced viral entry into host cells, and potentially with increasing infectivity and transmissibility (Bhattacharyya et al., 2020; Korber et al., 2020).

Additionally, we identified three substitutions in the nucleocapsid (N) gene of VEN-89312 changing GGG-to-AAC at positions 28,616-28,618 resulting R203K/G204R substitutions according to the whole-genome position after removing the 5’UTR. These R203K/G204R substitutions, which have been reported previously in other South American genomes, were absent from the two other Venezuelan genomes we sequenced. The alignment is shown highlighting the SNPs found in the N gene (Fig. 1D) (Crooks et al., 2004).

## 4. DISCUSSION

The presence of mutations previously reported to be associated with increased infectivity and transmissibility in the genomic sequences we determined for SARS-CoV-2 from both Venezuela and neighboring border regions of Colombia may represent an additional challenge for control of the COVID-19 pandemic in Latin America. The spectre of a SARS-CoV-2 variant with increased infectivity further complicates the already-complex epidemiological landscape with its chronic limitations of biomedical understaffing, poor healthcare infrastructure, limited or inadequate diagnostic capacities, and poor compliance of the general population with disease containment measures (Miller et al., 2020).

Following the first report of SARS-CoV-2 in Brazil in late February 2020 (Andrus et al., 2020; Rodriguez-Morales et al., 2020), SARS-CoV-2 has spread rapidly across the region, resulting in nearly two million cases as of June 25^th^, 2020, with Brazil and Peru reporting the highest burden of disease (“Epidemic Diseases - Cumulative suspected and confirmed COVID-19 cases reported by countries and territories in the Americas,” n.d.).

Although Colombia and Venezuela both implemented early lockdown and containment strategies since March 10^th^ and March 17^th^, respectively, (“CORONAVIRUS (COVID-19),” n.d., “Presidente Maduro anuncia que este martes todo el país entra en cuarentena social • Ministerio del Poder Popular para Relaciones Exteriores,” n.d.), according to current official records there have been over 124,500 COVID-19 cases in Colombia and over 7,700 cases in Venezuela (“CORONAVIRUS (COVID-19),” n.d., “Estadísticas Venezuela | COVID-19 en Venezuela,” n.d.). However, these numbers have to be contextualized because: (i) Colombia has exclusively performed state-of-the-art RT-PCR-based SARS-CoV-2 diagnosis whereas Venezuelan authorities have relied mostly on serological testing that are considerably less specific (Chia et al., 2020) and less useful than molecular methods for acute case detection; (ii) Venezuela has precarious epidemiological surveillance and contact tracing systems; and (iii) Venezuela has reduced international traffic through airports secondary to extensive departure of multiple airlines.

The concurrent humanitarian crisis has forced millions of Venezuelans to flee to neighboring countries ---mainly Colombia--- seeking economic and social stability (Torres and Castro, 2019; Tuite et al., 2018). Venezuelans with ongoing health issues also travel to Colombia to obtain high-quality healthcare such as surgical procedures and hemodialysis (Daniels, 2020). However, massive Venezuelan migration has resulted in an unprecedented infectious disease exodus, representing one of the most concerning public health threats in the region (Grillet et al., 2019; Torres and Castro, 2019). COVID-19 has deepened the situation and has prompted xenophobia and further marginalization of Venezuelan migrants and refugees at the Venezuelan-Colombian border (Daniels, 2020).

The shared presence of B lineages in SARS-CoV-2 from Venezuela and Colombia reinforces the close interactions of persons living in border regions and the difficulty of containment across a porous border. Similarly, the detection of the B.1.13 lineage, only previously described in cases in Spain, England and Australia (Batty et al., 2020) further underlines the rapid global spread of SARS-CoV-2 through interconnected populations. Additionally, the presence of substitution D614G in the spike protein of the three viruses from patients residing in the current hotspots of COVID-19 in Venezuela may correlate with the reported increased infectivity (Korber et al., 2020) observed in SARS-CoV-2-infected patients in the state of Zulia (“Zulia suma 15 muertes confirmadas por coronavirus y seis no reportadas - Efecto Cocuyo,” n.d.).

An important limitation to our study is that the limited number of genomes currently available from Venezuela could potentially result in sampling bias. Given the difficulty in obtaining samples, the extent to which our findings truly reflect Venezuela’s overall phylogenetic landscape remains to be determined. Future studies are needed to expand the SARS-CoV-2 genome repertoire in Venezuela and related areas, and to enable better understanding of the interplay between genotype and phenotype, and their relevance for disease surveillance and containment.

## 5. CONCLUSION

The Venezuelan humanitarian and refugee crisis, coupled with uncontrolled migration across the Colombian-Venezuelan border, is a devastating reminder of the potential effects of infectious disease spillover on the already vulnerable public health systems of neighboring countries. As SARS-CoV-2 continues to spread across Latin America, public health authorities and the international community should carefully follow the impact of the pandemic on displaced populations. Intensive efforts are urgently needed to help minimize the impact of Venezuela’s crisis on the COVID-19 pandemic crisis.

## Data Availability

All the data is available within the manuscript.

## 6. ACKNOWLEDGEMENTS

Funding was provided by the University of Glasgow, Scottish Funding Council and the Global Challenges Research Fund (GCRF) and GCRF Research Network EP/T003782/1.

